# Understanding end-user preferences for hand hygiene enabling technologies: a mixed-methods study in peri-urban Lusaka

**DOI:** 10.1101/2024.10.11.24315333

**Authors:** Katayi Kazimbaya, Katherine Davies, Mwamba M. Mwenge, Elisabeth Tadiri, Jenala Chipungu, Robert Dreibelbis

## Abstract

**Introduction:** Handwashing facilities (HWFs) are associated with higher rates of handwashing with soap, and the presence of a HWF is the global proxy indicator of household handwashing behaviour. There is limited information on attributes of HWFs important to end-users with few comparative assessments of HWFs from a user-perspective. We aimed to identify attributes of HWFs important to end-users and determine how pre-manufactured HWFs ranked against these attributes.

**Method:** We identified eight pre-existing HWFs: two locally manufactured (Kalingalinga bucket and Tippy tap) and six industrially produced designs (Jengu, SatoTap, SaniTap, HappyTap, SpaTap, and Kohler Cleanse). Two rounds of focus group discussions were conducted with a diverse group of targeted end-users in two peri-urban communities in Lusaka, Zambia. In the first, participants discussed aspects of each HWF they liked and disliked, and thematic analysis was used to define nine attributes for comparision across each HWF. In the second round, participants individually ranked each HWF against the identified attributes, their overall preference, and overall preference once estimated retail prices were revealed. Participants also ranked attributes by importance. Ranking data were modelled using rank-ordered logistic regression.

**Results:** Discussions revealed nine attributes important to end users: appearance, water management, hygienic use, convenience, water disposal, vulnerability to theft or breakage, ease of use, price and maintenance. Hygienic use and water management were considered most important attributes. Excluding price, facilities resembling a sink, such as the Happy Tap (34%) and Jengu (28%), had the highest probability of being ranked first. With consideration for price, participants preferred lower-cost HWFs such as the Kalingalinga bucket (44%), Tippy Tap (13%) and SATO Tap (10%).

**Conclusion:** This study identified nine attributes important to end-users that can inform future design efforts. Future work will explore user preferences *in situ* by rotating households through specific HWF for an extended period. Potential manufacturers should continue to iterate on HWF designs emphasizing on reducing costs.

## Introduction

Household access to a handwashing facility (HWF) with soap and water is crucial for achieving Sustainable Development Goal (SDG) 6.2, which aims to achieve adequate and equitable sanitation and hygiene for all by 2030 (United Nations 2015). The presence of an improved HWF in the home – defined as the presence of both soap and water in a single location for use in handwashing – is the global proxy indicator of handwashing behaviour (UNICEF, 2013). Handwashing with soap is effective at reducing the risk of infectious diseases, including diarrhoea and respiratory infection (Ross et al., 2023; Wolf et al., 2022). Global and regional disease outbreaks, such as COVID-19, cholera and typhoid, have magnified the need for improved hand hygiene practices (Berendes et al., 2022). However, global handwashing rates are low, particularly in low resource settings (Freeman et al., 2014; Wolf et al., 2019).

Access to a designated HWF is associated with higher rates of handwashing with soap (Luby et al., 2009; Wolf et al., 2019). Handwashing behaviour has also been attributed to physical characteristics of HWFs such as tap design and container size (Devine, 2010; Ezezika et al., 2023; Hulland et al., 2013). Limited access to handwashing infrastructure remains a key barrier to handwashing with soap in low-resource settings (Ezezika et al., 2023). In Zambia, over 80% of the population lack access to a handwashing facility with soap and water at home (WHO/UNICEF JMP 2022). To address this, several HWFs have been developed and piloted specifically targeting handwashing behaviours of end-users in resource limited settings (Biran, 2011; Coultas et al., 2020; Hulland et al., 2013; Husain et al., 2015; Revell & Huynh, 2018; SNV 2020; Whinnery et al., 2016).

There is limited peer-reviewed literature exploring the attributes of HWFs that potential end-users prioritise and few comparative assessments of multiple HWFs from a user perspective. While user preferences are frequently incorporated into the design process of specific HWFs, available studies focus primarily on a single improved HWF design compared to traditional facilities or focus on the physical performance of the HWF (Biran, 2011; Brial et al., 2023; Devine, 2010; Hulland et al., 2013; Whinnery et al., 2016). Comprehensive comparative assessments of multiple HWF designs have been limited to small samples, a limited number of technologies, and use of investigator-defined comparative categories (Brial et al., 2023). This two-phased study aimed to address this gap in the literature by identifying the attributes of HWFs considered important by a diverse set of end-users from low-income, peri-urban communities in Zambia (Phase 1) before determining how these end-users rank multiple existing HWFs against these locally-defined attributes (Phase 2).

## Methods

### Study Setting and Study Design

The study was conducted in two peri-urban communities, George and Matero, located in the Western part of Lusaka, Zambia. As the capital and largest city in Zambia, Lusaka’s water and sanitation infrastructure is inadequate for the current population of over two million as rapid population growth has outpaced investments in these essential services (Vonk, 2021). Conditions are worse in the city’s unplanned and informal settlements (peri-urban areas), where 60-70% of the population reside (Chiwele et al., 2022). George and Matero are located geographically next to each other and have a combined population of over 320,000 residents. These communities are characterised by densely packed, informal housing arrangements, where access to WASH remains a persistent challenge. Most residents access water via water kiosks managed by the local council. Water supply from the municipality is also largely provided through communal boreholes accessed during limited times in a day at a minimal fee. Flooding coupled with poor waste management often results in cyclical cholera outbreaks and other public health issues in these communities (Hubbard et al., 2020; Idoga et al., 2019). Matero is considered the more affluent community, with more households accessing water piped into their yard or plot. These communities were conveniently selected based on the current close partnership the Centre for Infectious Disease Research in Zambia (CIDRZ) holds with the local health facility in the communities.

Our study was completed in two phases – a qualitative exploratory phase used to define key attributes and a second quantitative phase in which potential end users ranked facilities according to each of the attributes identified. Below, we present the methods and results for each phase sequentially with a combined discussion.

### Ethical Considerations

Ethical approval was obtained from London School of Hygiene and Tropical Medicine, Research Ethics Committee, London, UK (Ref: 29745) and the University of Zambia Biomedical Research Ethics Committee (Ref: UNZABREC 4329-2023). All study staff were trained in Human Subject Protection (HSP). Prior to study activities, an information sheet explaining the study procedure and aims was read to participants. Informed written consent was obtained from all study participants in their native language by written signature or thumbprint, depending on literacy status.

## Phase 1: Attribute Determination

### Methods

#### Handwashing facilities

Following a scoping exercise, a total of eight HWFs were identified for use in the study: Tippy Tap, Kalingalinga bucket, SatoTap, SpaTap, Jengu, SaniTap, HappyTap and Kohler Cleanse (Figure 1) (Jengu 2024; Kohler 2024; mWater 2021; Revell & Huynh, 2018; SaniTap 2024; SNV 2020; SpaTap 2024). Selected facilities were those that were or could be manufactured locally (TippyTap, Kalinglinga Bucket) or were available from the manufacturer and could be shipped to the study site in Zambia.

**Figure 1.**
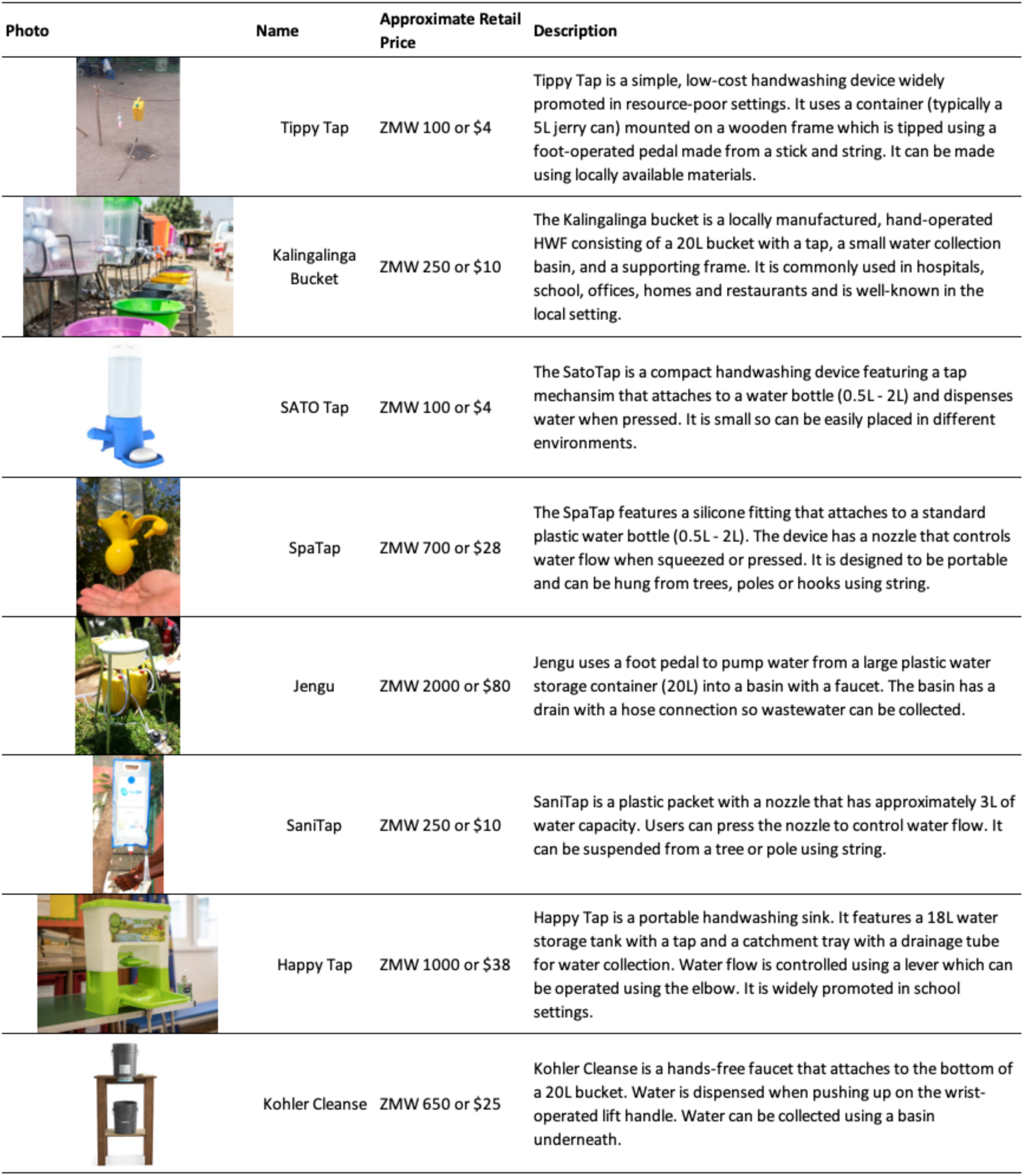
Summary of handwashing facilities included in the study.

#### Sampling Procedures

We randomly selected one community (George) from the two communities included for this phase of the study. A purposive sample of 37 individuals were selected across four participant groups: i) primary caregivers of under 5 children, ii) elderly populations (> 65 years old), iii) people living with disabilities and iv) adult men (age 18-64). Focus group discussions (FGDs) with 9-10 participants were held with each group separately. Due to the exploratory nature of this study, the number of focus groups was set to reflect anticipated diversity between key targeted groups of end-users. Community Health Workers (CHWs) familiar with the communities helped to identify participants from each group. Participants were approached by neighbourhood health committee members (NHCs) and invited to participate.

#### Data Collection

FGDs were completed in October 2023 in the community health clinic. A team of four research assistants facilitated the FGDs (two males, two females), all of whom held a Bachelor’s degree. Two research assistants were assigned per FGD (one as facilitator and one as note-taker). The team were managed throughout data collection by the lead author (KK, PhD). Research assistants were fluent in the local language (Nyanja) and had previous experience collecting data related to water, sanitation and hygiene. Prior to data collection, research assistants completed training on FGD facilitation, interview techniques, and ethical safeguarding (informed consent and data protection).

During the FGDs, participants were allocated five minutes to use each hand washing facility (HWF) unguided and thereafter the facilitator showed the participants how each HWF functioned. The participants were then asked to discuss each HWF as a group and reflect on which aspects of the HWFs they liked the most and least. FGDs were audio recorded and detailed notes were completed by research assistants. FGDs lasted around 90 minutes.

#### Data Analysis

Qualitative data were analysed according to the Braun and Clarke thematic analysis method (Braun & Clarke, 2006). Through thematic analysis of FGD transcriptions and detailed notes, the most salient attributes were identified and compared across users and across discussions of specific HWFs. This generated a list of emergent attributes which were further refined and defined through discussion by the study team.

### Results

A total of 37 individuals participated across four FGDs, nine from each participant interest group except for adult men which had 10 participants. Most participants lived on a shared plot, collected water from a public tap or standpipe and did not have a handwashing facility (Table S1). Thematic analysis of FGD transcripts identified 9 key attributes that reflected various components of user preference for HWF design. These were: appearance, water management, ease of use, hygienic use, convenience, water disposal, maintenance, vulnerability and price (Table 1).

**Table 1.** | Attribute determination and associated descriptions.

| Attribute | Definition | Example quote |
| --- | --- | --- |
| Appearance | How the HWF looks and how it would look in the home; how appealing the design is | *"We have said that E (Happy Tap) and H (Jengu) are good for the home because they look smart because the white color and the stand are nice, the containers and pipes are also nice, and when you put it in the house it makes it look smart." (P5-Participant with disability)*<br>*"The color is nice, and it makes the house look smart (Happy Tap)." (P3-Participant with disability)*<br>*"It also looks like a 'tit' (feeding bottle for a baby), a baby would start sucking it (Spa Tap)" (P1-Adult Men)* |
| Water management | Ease with which the HWF can be filled, how frequently it needs to be refilled, and how easy it is to determine how much water is available | *"There is need to increase the size of the water storage container because it cannot sustain you from morning to late afternoon (SATO Tap)." (P1-Caregiver)*<br>*"The same issue of removing it to refill and putting it back between the ropes/string becomes extra work plus the size also, you would have to refill about seven times per day (Spa Tap)." (P3-Adult Men)*<br>*"You cannot use it for the entire family because the water storage capacity is small. So, it can't sustain the entire family unless a few people, one or two (SATO Tap)." (P1-Adult Men)* |
| Ease of use | Ease with which hands can be washed by all members of the household (adults, children, disabled) | *"Maybe height for children should be considered when making the facility which could be slightly lower (Jengu)." (P3-Elderly)*<br>*"I have seen that there is labor on the foot not everyone can manage, holding and pressing with the foot like that (Jengu)." (P9-Participant with disability)* |
|  |  | *"The other thing making it difficult to use is the opener used to open the facility unless, if it was an auto sensor where you just put your hands and water starts flowing, it becomes easy that way (Kohler)." (P5-Adult Men)* |
| Hygienic Use | How hygienic it is to use the facility; if the facility is dirty after use; cross contamination avoided | *"The only challenge is with the tap when opening because, you will come with dirty hands and open it to wash your hands. However, how then do you close it? Because, as you close it, you will get back the dirt on your hands (Kalingalinga)." (P6-Adult Men)*<br>*"I also like it, it's nice because I can wash my hands without touching the container because of the foot peddle mechanism (Tippy Tap)." (P1-Caregiver)* |
| Convenience | Accessibility for all members of the households; ability to place the HWF at the location needed | *"(Kalingalinga) is also simple to be put in the dining room, because it can be used anywhere. It is good because you can even put it outside." (P6-Elderly)*<br>*"This idea is good but not for home use unless for programs like conferences and camps because it's easy to carry and easy to use. It is not needed for home use because its small and it has a plastic material which can get spoiled at any time (Sani Tap)." (P1-Adult Men)* |
| Water disposal | Ease of collecting, disposing, and/or reusing water after hand washing | *"I have seen it to be very helpful in the way we use it to wash hands because the dirty water does not go back in the container with clean water after handwashing (Jengu)." (P1-Elderly)*<br>*"I would prefer (Jengu) because it has a good water storage capacity and drainage system which prevents children from playing with dirty water compared to B (Tippy Tap) with a small container and children can be playing with dirty water in the dish." (P6-Adult Men)* |
| Maintenance | Ability to clean and maintain the facility, locate spare parts in case of breakage, cost of repairs | *"(Kalingalinga) is common because it is found in restaurants and other places. It is too common based on how much it costs. Its material is not too difficult to source, you could be passing by and have a metallic stand welded and buy a bucket to put on the stand and that will be all." (P9-Elderly)*<br>*"I would prefer B (Tippy Tap) because let's say the rope gets spoiled, I can easily look for another one to replace it as long as it is strong. Conversely, if the pump for H (Jengu) gets spoiled I would fail to know where to get it." (P4-Adult Men)* |
|  |  | *"Regarding water pressure, just in case you are in the rural areas where there is water with some dirty particles, those holes are too small and they can easily be blocked and the water would stop flowing. So, it can be a difficult facility to use if the water we are using has some dirty particles (Kohler)." (P1-Adult Men)* |
| Vulnerability | Likelihood of theft, loss, or damage | *"Children will remove the yellow thing in front of the bottle thinking it's a toy and they can spoil it (Spa Tap)." (P2-Caregiver)*<br>*"The mirror is attractive, and this is not good for the public because, it can easily break or be stolen (Jengu)." (P10-Adult Men)* |
| Price | How expensive is the facility | *"This one is good, affordable and it is also cheap. Wherever you go, whether it's in schools or clinics they manage to buy it. We also manage to buy it even in our homes (Kalingalinga)." (P3-Caregiver)*<br>*"B (Tippy Tap) is good because even those that cannot afford to have the stand made can just use the 'Y' trees and make it." (P4-Caregiver)*<br>*"Yes we have said they (H -Jengu and E- Happy Tap) are nice and make the house look smart but they are expensive. We would want to have them however what are we going to use to buy them we do not have money." (P8-Participant with disability).* |

## Phase 2: Ranking of HWFs

### Methods

#### Sampling Procedures

A total of 8 focus group discussions (FGDs) were conducted in George and Matero with four groups of participants (one in each community): i) primary caregivers of under 5 children, ii) elderly populations (> 65 years old), iii) adult women (age 18-64) and iv) adult men (age 18-64). Eligiblity was limited to only those participants who did not particiate in the first round of data collection. In addition to the new participants, two additional FGDs – one with the adult men and one with the caregivers of children under the age of five - that took part in Phase 1 were invited back to participate in the ranking to assess if involvement in previous phases of the study impacted rankings. Focus group discussions were held with each group separately. Each FGD had between eight and nine participants. Due to the exploratory nature of this study, the number of focus groups was set to reflect anticipated diversity between key targeted groups of end-users. Community Health Workers (CHWs) familiar with the communities helped to identify participants from each group. Participants were approached by NHCs in communities and invited to participate.

#### Data Collection

FGDs were completed between 15 January to 08 Februrary 2024 in community health clinics. A team of four research assistants facilitated the FGDs (two males, two females), all of whom held a Bachelor’s degree. The team were managed throughout data collection by the lead author (KK, PhD). Research assistants were fluent in the local language and had previous experience collecting data related to water, sanitation and hygiene. Prior to data collection, research assistants completed training on FGD facilitation, interview techniques, and ethical safeguarding (informed consent and data protection).

The participants were allocated five minutes to use each facility unguided and thereafter the facilitator showed the participants how each handwashing facility functioned. No information on the retail price of the facilities was shared to limit the extent to which perceived affordability would limit preferences. Participants were given laminated cards with images of each facility. Attributes from phase 1 (excluding price) were introduced one at a time and participants asked to individually rank each HWF from best to worst for each attribute. (). Ties were permitted. Participants were then asked to rank each HWF by their overall preference from most to lease desirable. Estimated retail prices (based on cost price from manufacturer or distributor as well as shipping for HWFS manufactured outside Zambia) were then shared and participants were asked to again rank by overall preference. Finally, participants were asked to rank each of the attributes in Phase 1 (Table 1) from most to least important.

#### Data Analysis

Ranking data on handwashing facility preferences and attribute importance was modelled using rank-ordered logistic regression by maximum likelihood using the *rologit* command in STATA 18 (StataCorp, College Station, TX). Models estimate the probability that a handwashing facility would be ranked first by a respondent against each attribute of interest and estimate the probability that an attribute would be ranked as the most important. Rank-ordered logistic regression considers all ranks assigned to an item, unlike conditional logit models, so two items with equal numbers of first place rankings can be differentiated using the rank-ordered model by how many lower rankings they received. To test if rankings varied between participants from different study sites (George and Matero) and participant groups (adult men, adult women, elderly (65+) and caregivers), rank-ordered models with interaction terms were fitted and Wald tests were performed. Variation in rankings between participants returning from phase 1 versus participants new to the study was also explored. Otherwise, ranking data from all participant groups were combined. Separate models were fitted to estimate predicted probabilities amongst participants from different study sites and participant groups.

### Results

A total of 81 individuals participated across ten focus group discussions (Table 2). Sixty percent (49/81) of participants were female and most had at least a primary education (Table S2). Participants mostly lived on a shared plot and did not own a handwashing facility. Participants from Matero were more likely to have received an education beyond primary-level, have formal employment and have water piped into their compound.

**Table 2.** | Participant Numbers.

| FGD Number | Community | Group | No of Participants | Returning from Phase 1 |
| --- | --- | --- | --- | --- |
| 1 | George | Adult Men | 8 | No |
| 2 | George | Adult Men | 8 | Yes |
| 3 | George | Adult Women | 9 | No |
| 4 | George | Caregivers | 8 | No |
| 5 | George | Caregivers | 8 | Yes |
| 6 | George | Elderly | 8 | No |
| 7 | Matero | Adult Men | 8 | No |
| 8 | Matero | Adult Women | 8 | No |
| 9 | Matero | Caregivers | 8 | No |
| 10 | Matero | Elderly | 8 | No |

#### Handwashing Facility Rankings

The Kalingalinga bucket had the highest probability of being ranked first across most attributes, including water management (47%), ease of use (41%), maintenance (37%), water disposal (32%), vulnerability (least vulnerable) (27%) and convenience (26%) (Table 3). Happy Tap and Jengu also performed well, ranking highly across most attributes. Handwashing facilities which participants could use without touching, such as the Happy Tap (35%), Jengu (33%) and Tippy Tap (11%), ranked highly for hygienic use. Furthermore, larger facilities, such as Kalingalinga (47%), Happy Tap (22%) and Jengu (15%) ranked highly for water management. Locally available handwashing facilities, such as the Kalingalinga bucket (37%) and the Tippy Tap (24%), ranked highly for maintenance while handwashing facilities new to the local context, such as the Happy Tap (8%) and Sanitap (6%), were considered vulnerable. We observed some variation in rankings by participant group (Figure S1).

**Table 3.** | Probability that each handwashing facility is ranked first against each attribute according to rank ordered logistic regression. *Stars represent where p-values fell when comparing each item’s log odds of being ranked higher compared to a reference item (ref)*.

| HWF | Probability HWF Ranked First (%) |  |  |  |  |  |  |  |  |  |  |  |  |  |  |  |
| --- | --- | --- | --- | --- | --- | --- | --- | --- | --- | --- | --- | --- | --- | --- | --- | --- |
|  | Appearance |  | Water Management |  | Ease of Use |  | Hygienic Use |  | Convenience |  | Water Disposal |  | Maintenance |  | Vulnerability (least vulnerable) |  |
| Happy Tap | 50% | *** | 22% | *** | 25% | *** | 35% | *** | 15% | * | 18% | *** | 3% | *** | 7% |  |
| Jengu | 19% | *** | 15% | *** | 16% | *** | 33% | *** | 8% |  | 19% | *** | 5% | ** | 17% | *** |
| Kalingalinga | 15% | *** | 47% | *** | 41% | *** | 8% | *** | 26% | *** | 31% | *** | 37% | *** | 27% | *** |
| Kohler | 4% | ** | 9% | *** | 2% | * | 3% |  | 10% |  | 14% | *** | 6% |  | 16% | *** |
| Sanitap | 3% |  | 2% | *** | 3% |  | 3% |  | 11% |  | 5% | * | 8% |  | 6% |  |
| Satotap | 3% |  | 1% | * | 4% | * | 5% | ** | 15% | * | 4% |  | 9% |  | 6% |  |
| Tippytap | 4% | * | 3% | *** | 5% | ** | 11% | *** | 5% | *** | 6% | ** | 24% | *** | 15% | *** |
| Spatap | 3% | (Ref) | 1% | (Ref) | 3% | (Ref) | 3% | (Ref) | 10% | (Ref) | 3% | (Ref) | 8% | (Ref) | 6% | (Ref) |

\*P<0.05
\*\*P<0.01
\*\*\*P<0.001

Without consideration for price, the Happy Tap had the highest probability of being ranked first overall (34%) followed by Jengu (28%) and Kalingalinga (24%) (Figure 2). However, when participants were informed of the retail price of each handwashing facility, the probability of Happy Tap being ranked first decreased to 9% and the probability of Jengu being ranked first decreased to 5%. With consideration for pricing, participants were most likely to rank Kalingalinga first (44%) followed by Tippy tap (13%) and Satotap (10%) (Figure 2) (Table S3).

**Figure 2.**
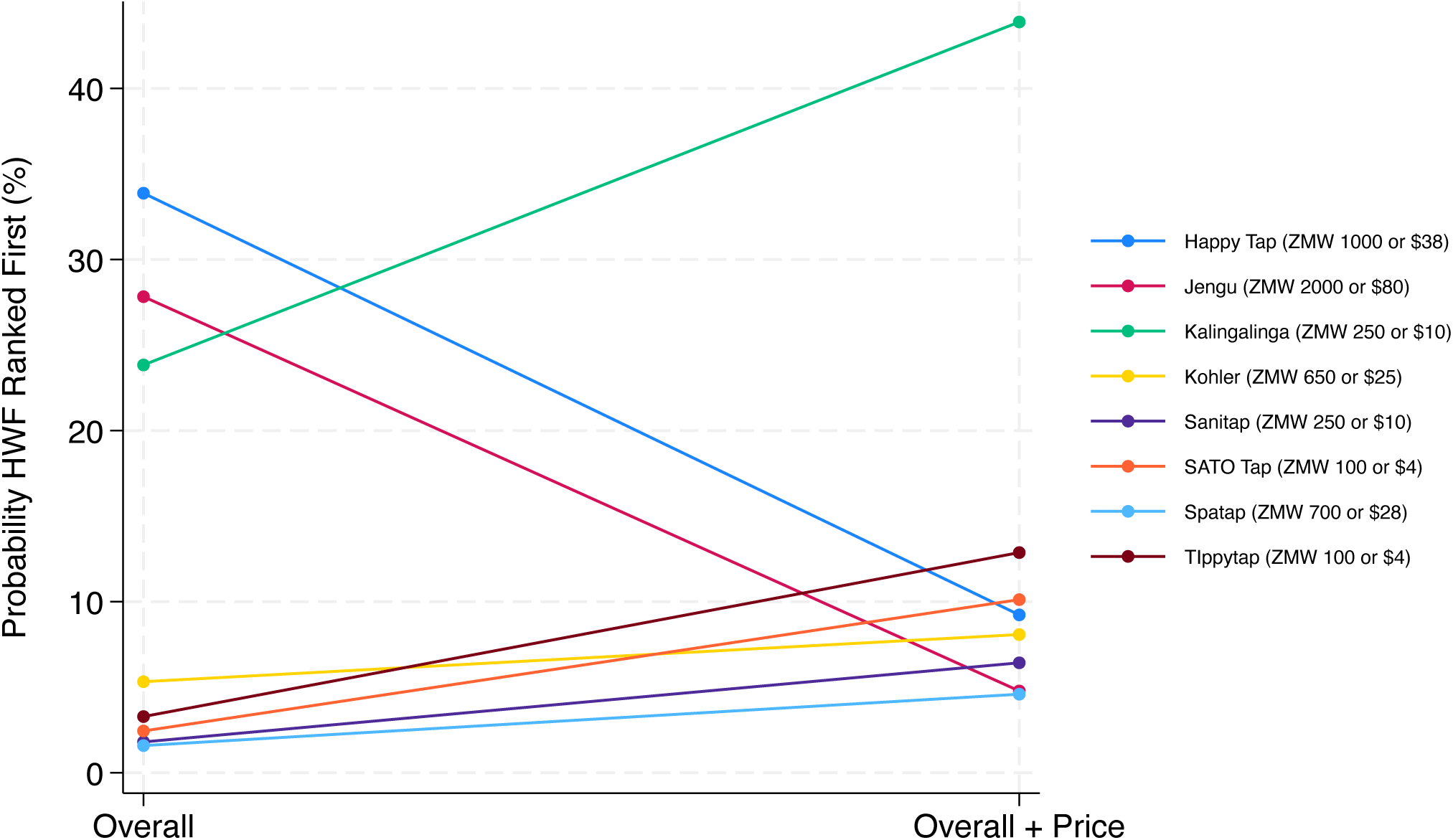
Probability of each handwashing facility being ranked first overall before and after consideration for price. *Retail prices provided to participants are shown in the key*.

Wald tests found variance in overall HWF rankings between different participant groups before (P<0.001) and after (P=0.018) consideration for price (Figure 3). Happy Tap, Jengu and Kalingalinga ranked highly amongst all participant groups before price was considered (Figure 3A) (Table S4). Adult women and caregivers were more likely to rank Jengu first, while adult men and elderly participants were more likely to rank the Kalingalinga bucket first. With consideration for price, participants from all groups ranked Kalingalinga the highest, with limited variation in rankings for lower ranking handwashing facilities noted between participants from different groups (Figure 3B) (Table S5). Limited variation in HWF rankings was found between different study sites (Figure S2) (Table S6) (Table S7). While there was some variance in overall rankings with consideration for price between participants returning from Phase 1 vs new to the study (P=0.007), the removal of participants from Phase 1 made minimal difference to the results (Table S8) (Table S9).

**Figure 3.**
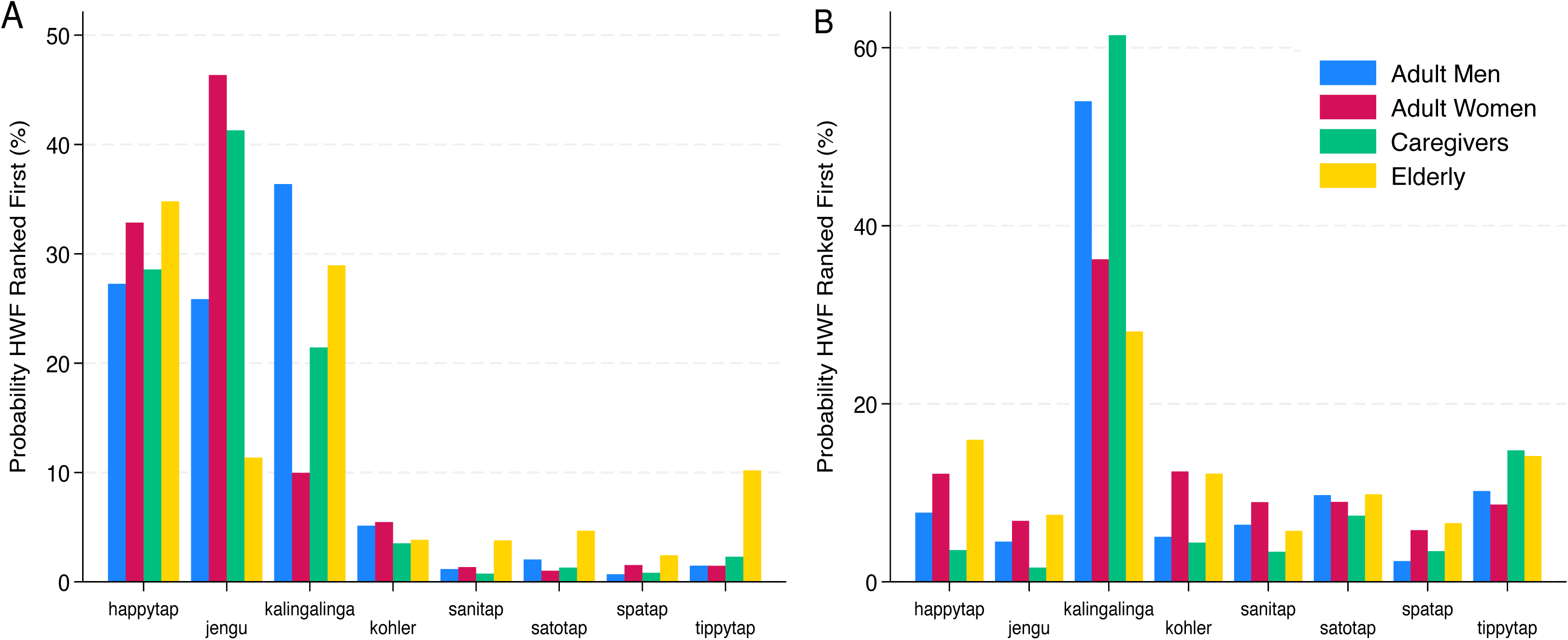
Probability of each handwashing facility being ranked first overall before (A) and after (B) consideration for price, stratified by group. *Predicted probabilities are estimated using rank-ordered logistic regression. Wald tests found variance in overall HWF rankings before (P<0.001) and after (P=0.018) consideration for price*.

#### Attribute Rankings

Hygienic use (19%) and water management (18%) were considered the most important attributes, while vulnerability (6%) was considered the least important attribute amongst all participants (Table 4). Wald tests found the ranked importance of attributes varied between participant groups (P=0.02) and study sites (P<0.001), and between participants returning versus new to the study (P=0.002). Appearance was considered important amongst elderly participants (17%) and adult women (16%) but was not considered as important by adult men (9%) and caregivers (7%). Price was considered the most important attribute amongst caregivers (20%) but was not ranked highly amongst other groups. Participants from Matero were highly likely to rank price as the most important attribute (17%), while participants from George were not (5%).

**Table 4.**
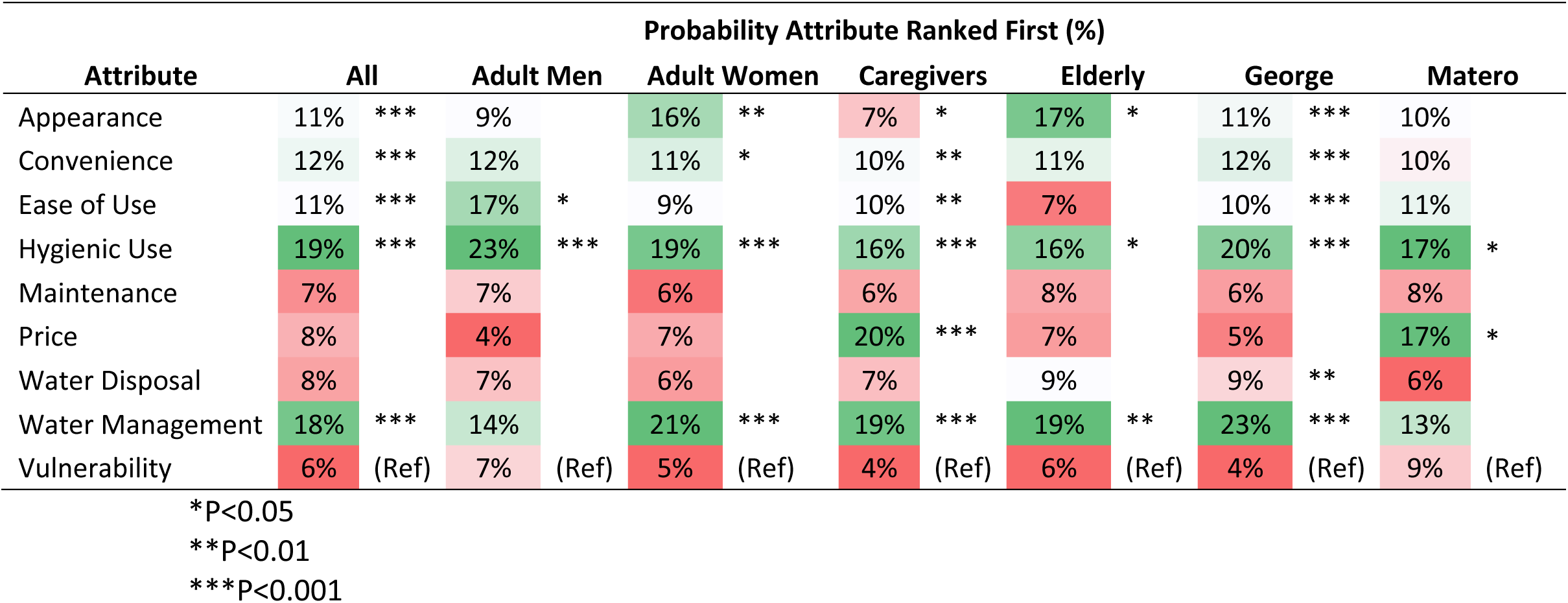
| Probability of each attribute being ranked as most important. Stars represent where p-values fell when comparing each item’s log odds of being ranked higher compared to a reference item (ref).

## Discussion

Presence of an improved HWF with both soap and water is associated with improved handwashing behaviour at key moments (Wolf et al., 2019). We identified a range of locally and globally manufactured designs with the potential to facilitate HWWS in the domestic environment, ranging from simple devices that are usable with widely available plastic bottles to devices that function as a kitchen sink. By comparing these multiple designs, our study has identified a core set of attributes important to a diverse group of end-users that can be used to inform HWF design: ease of use, water management, convenience, appearance, hygiene use, water disposal, maintenance, vulnerability and price. Hygienic use and water management were considered the most important attributes, and vulnerability the least important attribute. Without consideration for price, facilities which resembled a sink, such as the Happy Tap and Jengu, had the highest probability of being ranked first overall. With consideration for price, participants instead preferred lower-cost HWFs such as the Kalingalinga bucket, Tippy Tap and SATO Tap.

Caregivers – who are usually responsible for purchasing decisions related to the domestic environment in this setting – ranked price as the most important attribute. However, other groups instead ranked attributes such as “hygienic use” (adult men) or “appearance” (adult women and elderly) higher than caregivers. Adult men in this setting typically work in jobs where their hands get dirty, which could explain their preference for a facility which is easy and hygienic to use. Price was considered more important in Matero, the more affluent community. While unexpected, this could be due to the inclusion of additional participants returning from Phase 1 in George, who ranked price as one of the least important attributes (Table S10). Despite this, the change in HWF rankings following the revelation of cost data illustrates the importance of retail price in driving end-user preferences. The literature identifies cost and affordability as a key barrier to handwashing with water and soap globally (Ezezika et al., 2023; Kisaakye et al., 2020). Therefore, it is crucial manufacturers consider the cost of HWFs in the design process.

Our findings are consistent with a comparative assessment of seven HWFs conducted in Tanzania, which found the Happy Tap was the favoured facility without consideration for price (Brial et al., 2023). This study found that end-users liked the Happy Tap due to its attractive and modern characteristics, aligning with the high rankings Happy Tap received for appearance in our study. Several studies found end-users aspire to own facilities with a high aesthetic value, and the appearance of facilities is important for use (Devine, 2010; Hulland et al., 2013; Revell & Huynh, 2018). In this study, appearance was considered important by adult women and elderly participants.

Hygienic use was considered the most important attribute to participants. Hands-free handwashing facility designs, such as Happy Tap, Jengu and Tippy Tap were ranked highly for hygienic use. These findings align with a case study in Uganda which found end-users preferred the Tippy Tap over standard jerry cans as the foot-pedal helped to avoid contamination (Biran, 2011). Foot pedal-based systems have increased in popularity following the COVID-19 pandemic, in an effort to limit contamination (SNV 2020). A scoping review of hand hygiene guidelines found guidelines commonly recommend COVID-19-related adaptations to hand hygiene stations to limit cross contamination (MacLeod et al., 2023). Therefore, HWF designs that limit cross-contamination should be prioritised.

Water management was also ranked as one of the most important attributes by end-users. HWFs with a large water storage capacity, such as the Kalingalinga bucket and Happy Tap, ranked highly for water management while HWFs with a small water storage capacity, such as the SATO Tap and SaniTap ranked poorly. Previous studies exploring end-user preferences highlight the Happy Tap was favoured due to its integrated capacity for storing water while the SpaTap was one of the least popular solutions due to its small water storage capacity (Brial et al., 2023). Water storage capacity of HWFs is important for acceptability and facilities requiring frequent refilling are not conducive to repeated use throughout the day (Biswas et al., 2017; Devine, 2010; Ezezika et al., 2023; Hulland et al., 2013). Most participants in this study do not have water piped into their dwelling. Water storage capacity was ranked as particularly important amongst women and caregivers who are generally responsible for water collection. With climate change expected to exacerbate the burden of water collection on women’s welfare, water storage capacity of handwashing facilities will remain an important attribute of HWF designs to end-users (Carr et al., 2024).

We acknowledge the limitations of this study. First, participants were only given a limited amount of time to use each of the HWFs in controlled settings before being asked to rank them. Therefore, this study does not consider how participants would rank the HWFs after using them for a longer period in a household setting. This limitation will be addressed in a subsequent Trial of Improved Practices (TIPS) with top-ranking handwashing facilities. Secondly, a Hausman test revealed the same decision weights were not applied with higher and lower ranking HWFs, with rank-ordering of lower ranking HWFs more random than higher ranking HWFs (Hausman & Ruud, 1987). Therefore, the regression model’s ability to predict HWF rankings is less predictable for lower-ranked items. Thirdly, it is possible that comments from other participants could have biased participant rankings through social desirability mechanisms. However, this was mitigated by conducting rankings individually. Fourthly, purposive sampling techniques used in this study mean results may not be generalisable beyond populations with similar characteristics to those included in our study. Finally, it is important to note the results of this study may not be applicable to different settings, such as emergencies, where different attributes of HWFs are likely to be prioritised (Husain et al., 2015).

## Conclusion

This study identified a set of HWF attributes that were important to a diverse set of end-users which can be used to inform future design efforts. Hygienic use and water management were considered the most important attributes, and vulnerability the least important attribute. In the absence of cost data, HWFs that resembled a traditional sink, such as the Happy Tap and Jengu, were the highest ranking HWFs. Price further informed user preferences, with participants instead preferring lower cost HWFs such as the Kalingalinga bucket, Tippy Tap and SATO Tap. Potential manufacturers should continue to iterate on HWF design with an emphasis on reducing costs. Future work will explore user preferences in situ by rotating households through specific HWFs for an extended period.

## Data Availability

All data produced in the present study are available upon reasonable request to the authors.

## References

Berendes, D., Martinsen, A., Lozier, M., Rajasingham, A., Medley, A., Osborne, T., Trinies, V., Schweitzer, R., Prentice-Mott, G., Pratt, C., Murphy, J., Craig, C., Lamorde, M., Kesande, M., Tusabe, F., Mwaki, A., Eleveld, A., Odhiambo, A., Ngere, I.,… Handzel, T. (2022). Improving water, sanitation, and hygiene (WASH), with a focus on hand hygiene, globally for community mitigation of COVID-19. PLOS Water, 1(6), e0000027. 10.1371/journal.pwat.0000027

Biran, A. (2011). Enabling technologies for handwashing with soap : a case study on the tippy-tap in Uganda (English). Water and sanitation program working paper. W. B. Group. http://documents.worldbank.org/curated/en/805931468117886154/Enabling-technologies-for-handwashing-with-soap-a-case-study-on-the-tippy-tap-in-Uganda

Biswas, D., Nizame, F. A., Sanghvi, T., Roy, S., Luby, S. P., & Unicomb, L. E. (2017). Provision versus promotion to develop a handwashing station: the effect on desired handwashing behavior. BMC Public Health, 17(1), 390. 10.1186/s12889-017-4316-6

Braun, V., & Clarke, V. (2006). Using thematic analysis in psychology. Qualitative Research in Psychology, 3, 77–101. 10.1191/1478088706qp063oa

Brial, E., Aunger, R., Muangi, W. C., & Baxter, W. (2023). Development of a novel hand cleansing product for low-income contexts: The case of tab soap. PLOS ONE, 18(5), e0283741. 10.1371/journal.pone.0283741

Carr, R., Kotz, M., Pichler, P.-P., Weisz, H., Belmin, C., & Wenz, L. (2024). Climate change to exacerbate the burden of water collection on women’s welfare globally. Nature Climate Change. 10.1038/s41558-024-02037-8

Chiwele, D., Lamson-Hall, P., & Wani, S. (2022). “Informal Settlements in Lusaka”. International Growth Centre and the UN-Habitat.

Coultas, M., Iyer, R., & Myers, J. (2020). Handwashing Compendium for Low Resource Settings: A Living Document, Edition 1, The Sanitation Learning Hub, Brighton: IDS. IDS.

Devine, J. (2010). Beyond tippy-taps: The role of enabling products in scaling up and sustaining handwashing. Waterlines, 29(4), 304–314. http://www.jstor.org/stable/24686636

Ezezika, O., Heng, J., Fatima, K., Mohamed, A., & Barrett, K. (2023). What are the barriers and facilitators to community handwashing with water and soap? A systematic review. PLOS Global Public Health, 3(4), e0001720. 10.1371/journal.pgph.0001720

Freeman, M. C., Stocks, M. E., Cumming, O., Jeandron, A., Higgins, J. P. T., Wolf, J., Prüss-Ustün, A., Bonjour, S., Hunter, P. R., Fewtrell, L., & Curtis, V. (2014). Systematic review: Hygiene and health: systematic review of handwashing practices worldwide and update of health effects. Tropical Medicine & International Health, 19(8), 906–916. 10.1111/tmi.12339

Hausman, J. A., & Ruud, P. A. (1987). Specifying and testing econometric models for rank-ordered data. Journal of Econometrics, 34(1), 83–104. 10.1016/0304-4076(87)90068-6

Hubbard, S. C., Meltzer, M. I., Kim, S., Malambo, W., Thornton, A. T., Shankar, M. B., Adhikari, B. B., Jeon, S., Bampoe, V. D., Cunningham, L. C., Murphy, J. L., Derado, G., Mintz, E. D., Mwale, F. K., Chizema-Kawesha, E., & Brunkard, J. M. (2020). Household illness and associated water and sanitation factors in peri-urban Lusaka, Zambia, 2016–2017. npj Clean Water, 3(1), 26. 10.1038/s41545-020-0076-4

Hulland, K. R. S., Leontsini, E., Dreibelbis, R., Unicomb, L., Afroz, A., Dutta, N. C., Nizame, F. A., Luby, S. P., Ram, P. K., & Winch, P. J. (2013). Designing a handwashing station for infrastructure-restricted communities in Bangladesh using the integrated behavioural model for water, sanitation and hygiene interventions (IBM-WASH). BMC Public Health, 13(1), 877. 10.1186/1471-2458-13-877

Husain, F., Hardy, C., Zekele, L., Clatworthy, D., Blanton, C., & Handzel, T. (2015). A pilot study of a portable hand washing station for recently displaced refugees during an acute emergency in Benishangul-Gumuz Regional State, Ethiopia. Conflict and Health, 9(1), 26. 10.1186/s13031-015-0053-6

Idoga, P. E., Toycan, M., & Zayyad, M. A. (2019). Analysis of Factors Contributing to the Spread of Cholera in Developing Countries. Eurasian J Med, 51(2), 121–127. 10.5152/eurasianjmed.2019.18334

Jengu (2024). Jengu Handwashing Unit. Retrieved 24 June 2024 from https://jengu.org.uk/

Kisaakye, P., Ndagurwa, P., & Mushomi, J. (2020). An assessment of availability of handwashing facilities in households from four East African countries. Journal of Water, Sanitation and Hygiene for Development, 11(1), 75–90. 10.2166/washdev.2020.129

Kohler (2024). Kohler Cleanse. Retrieved 24 June 2024 from https://www.kohlercompany.com/social-impact/innovation-for-good/ifg-products/cleanse/

Luby, S. P., Halder, A. K., Tronchet, C., Akhter, S., Bhuiya, A., & Johnston, R. B. (2009). Household characteristics associated with handwashing with soap in rural Bangladesh. Am J Trop Med Hyg, 81(5), 882–887. 10.4269/ajtmh.2009.09-0031

MacLeod, C., Braun, L., Caruso, B. A., Chase, C., Chidziwisano, K., Chipungu, J., Dreibelbis, R., Ejemot-Nwadiaro, R., Gordon, B., Esteves Mills, J., & Cumming, O. (2023). Recommendations for hand hygiene in community settings: a scoping review of current international guidelines. BMJ Open, 13(6), e068887. 10.1136/bmjopen-2022-068887

mWater (2021). SATO Tap rapid evolution report. https://sato.lixil.com/wp-content/uploads/2023/02/SATO-TAP-trial-report-Africa.pdf

Revell, G., & Huynh, N. A. (2018). HappyTap: aspirational handwashing device commercialization in Vietnam https://repository.lboro.ac.uk/articles/conference_contribution/HappyTap_aspirational_handwashing_device_commercialization_in_Vietnam/9595961

Ross, I., Bick, S., Ayieko, P., Dreibelbis, R., Wolf, J., Freeman, M. C., Allen, E., Brauer, M., & Cumming, O. (2023). Effectiveness of handwashing with soap for preventing acute respiratory infections in low-income and middle-income countries: a systematic review and meta-analysis. Lancet, 401(10389), 1681–1690. 10.1016/s0140-6736(23)00021-1

SaniTap (2024). SaniTap. Retrieved 24 June 2024 from https://sanitap.org/

SNV (2020). Practical options for handwashing stations: A guide for promoters and producers, Technical Paper, The Hague. https://a.storyblok.com/f/191310/0ae1aa224e/202012-practical-_options-for-handwashing-stations.pdf

SpaTap (2024). SpaTap. Retrieved 24 June 2024 from https://www.spatap.com/

UNICEF. (2013). Handwashing Promotion: Monitoring and Evaluation Module. https://www.unicef.org/media/91326/file/Handwashing-MandE-Module.pdf

United Nations (2015). Transforming our world: the 2030 Agenda for Sustainable Development. Retrieved 24 June 2024 from https://sdgs.un.org/2030agenda

Vonk, J. (2021). Sustainable Water and Sanitation in Zambia: Impact evaluation of the ‘Urban WASH’ project.

Whinnery, J., Penakalapati, G., Steinacher, R., Wilson, N., Null, C., & Pickering, A. J. (2016). Handwashing With a Water-Efficient Tap and Low-Cost Foaming Soap: The Povu Poa "Cool Foam" System in Kenya. Glob Health Sci Pract, 4(2), 336–341. 10.9745/ghsp-d-16-00022

WHO/UNICEF JMP (2022). Hygiene. Retrieved 24 June 2024 from https://washdata.org/data/household#!/table?geo0=country&geo1=ZMB

Wolf, J., Hubbard, S., Brauer, M., Ambelu, A., Arnold, B. F., Bain, R., Bauza, V., Brown, J., Caruso, B. A., Clasen, T., Colford, J. M., Jr., Freeman, M. C., Gordon, B., Johnston, R. B., Mertens, A., Prüss-Ustün, A., Ross, I., Stanaway, J., Zhao, J. T.,… Boisson, S. (2022). Effectiveness of interventions to improve drinking water, sanitation, and handwashing with soap on risk of diarrhoeal disease in children in low-income and middle-income settings: a systematic review and meta-analysis. The Lancet, 400(10345), 48–59. 10.1016/S0140-6736(22)00937-0

Wolf, J., Johnston, R., Freeman, M. C., Ram, P. K., Slaymaker, T., Laurenz, E., & Prüss-Ustün, A. (2019). Handwashing with soap after potential faecal contact: global, regional and country estimates. Int J Epidemiol, 48(4), 1204–1218. 10.1093/ije/dyy253

