## Supplementary Materials for "Understanding end-user preferences for hand hygiene enabling technologies: a mixed-methods study in peri-urban Lusaka"

**Table S1 | Characteristics of FGD participants**

| Demographic | | Adult Men (n=10) | Caregivers of under 5s (n=9) | People with disabilities (n=9) | Elderly (n=9) |
|---|---|---|---|---|---|
| | | **Mean (SD)** | **Mean (SD)** | **Mean (SD)** | **Mean (SD)** |
| Age (years) | | 27.70 (8.12) | 29.33 (8.56) | 56.56 (16.21) | 72.56 (6.62) |
| | | **N (%)** | **N (%)** | **N (%)** | **N (%)** |
| Gender | Male | 10 (100%) | 0 (0%) | 4 (44%) | 4 (44%) |
| | Female | 0 (0%) | 9 (100%) | 5 (56%) | 5 (55%) |
| Employment Status | Formal employee | 0 (0%) | 1 (11%) | 0 (0%) | 0 (0%) |
| | Casual work/Self-employed | 7 (70%) | 4 (44%) | 3 (33%) | 2 (22%) |
| | Housewife | 0 (0%) | 4 (44%) | 0 (0%) | 0 (0%) |
| | Unemployed/Retired | 3 (30%) | 0 (0%) | 6 (67%) | 7 (78%) |
| Education* | Up to Primary | 4 (44%) | 6 (67%) | 7 (78%) | 8 (89%) |
| | Secondary + | 5 (56%) | 3 (33%) | 2 (22%) | 1 (11%) |
| Shared Plot† | Yes | 9 (90%) | 7 (78%) | 6 (67%) | 3 (38%) |
| | No | 1 (10%) | 2 (22%) | 3 (33%) | 5 (63%) |
| Water piped into compound | Yes | 4 (40%) | 4 (44%) | 1 (11%) | 3 (33%) |
| | No | 6 (60%) | 5 (56%) | 8 (89%) | 6 (67%) |
| Handwashing facility | Yes | 0 (0%) | 0 (0%) | 1 (11%) | 3 (33%) |
| | No | 10 (100%) | 9 (100%) | 8 (89%) | 6 (67%) |

*N=9 for adult men

† N=8 for elderly participants

**Table S2 | Characteristics of FGD participants**

| Demographic | | George | | | | | Matero | | | | |
|---|---|---|---|---|---|---|---|---|---|---|---|
| | | Adult Men (n=16) | Adult Women (n=9) | Caregivers (n=16) | Elderly (n=8) | Total (n=49) | Adult Men (n=8) | Adult Women (n=8) | Caregivers (n=8) | Elderly (n=8) | Total (n=32) |
| | | Mean (SD) | Mean (SD) | Mean (SD) | Mean (SD) | | Mean (SD) | Mean (SD) | Mean (SD) | Mean (SD) | Mean (SD) |
| Age (years) | | 28.56 (6.41) | 28.33 (8.99) | 32.19 (9.06) | 67.38 (4.00) | 36.04 (15.89) | 34.25 (10.50) | 35.88 (11.76) | 28 (4.66) | 69.88 (2.70) | 42 (18.41) |
| | | N (%) | N (%) | N (%) | N (%) | | N (%) | N (%) | N (%) | N (%) | N (%) |
| Gender | Male | 16 (100%) | 0 (0%) | 0 (0%) | 4 (50%) | 20 (41%) | 8 (100%) | 0 (0%) | 0 (0%) | 4 (50%) | 12 (38%) |
| | Female | 0 (0%) | 9 (100%) | 16 (100%) | 4 (50%) | 29 (59%) | 0 (0%) | 8 (100%) | 8 (100%) | 4 (50%) | 20 (63%) |
| Employment Status* | Formal employee | 1 (6%) | 0 (0%) | 1 (6%) | 0 (0%) | 2 (4%) | 4 (57%) | 3 (38%) | 0 (0%) | 0 (0%) | 7 (23%) |
| | Casual worker | 11 (69%) | 2 (22%) | 8 (50%) | 2 (25%) | 23 (47%) | 0 (0%) | 3 (38%) | 4 (50%) | 2 (25%) | 9 (29%) |
| | Housewife | 0 (0%) | 1 (11%) | 4 (25%) | 1 (13%) | 6 (12%) | 0 (0%) | 0 (0%) | 2 (25%) | 1 (13%) | 3 (10%) |
| | Unemployed/Retired | 4 (25%) | 6 (67%) | 3 (19%) | 5 (63%) | 18 (37%) | 3 (43%) | 2 (25%) | 2 (25%) | 5 (63%) | 12 (39%) |
| Education | Up to Primary | 8 (50%) | 3 (33%) | 9 (56%) | 6 (75%) | 26 (53%) | 1 (13%) | 2 (25%) | 4 (50%) | 5 (63%) | 12 (38%) |
| | Secondary + | 8 (50%) | 6 (67%) | 7 (44%) | 2 (25%) | 23 (47%) | 7 (88%) | 6 (75%) | 4 (50%) | 3 (38%) | 20 (63%) |
| Shared Plot** | Yes | 12 (75%) | 7 (78%) | 11 (69%) | 2 (29%) | 32 (67%) | 6 (75%) | 8 (100%) | 6 (75%) | 4 (50%) | 24 (75%) |
| | No | 4 (25%) | 2 (22%) | 5 (31%) | 5 (71%) | 16 (33%) | 2 (25%) | 0 (0%) | 2 (25%) | 4 (50%) | 8 (25%) |
| Water piped into compound | Yes | 9 (56%) | 5 (56%) | 9 (56%) | 2 (25%) | 25 (51%) | 6 (75%) | 7 (88%) | 6 (75%) | 6 (75%) | 25 (78%) |
| | No | 7 (44%) | 4 (44%) | 7 (44%) | 6 (75%) | 24 (49%) | 2 (25%) | 1 (13%) | 2 (25%) | 2 (25%) | 7 (22%) |
| Handwashing facility* | Yes | 5 (31%) | 4 (44%) | 3 (19%) | 5 (53%) | 17 (35%) | 2 (29%) | 2 (25%) | 6 (75%) | 4 (50%) | 14 (45%) |
| | No | 11 (69%) | 5 (56%) | 13 (81%) | 3 (38%) | 32 (65%) | 5 (71%) | 6 (75%) | 2 (25%) | 4 (50%) | 17 (55%) |

*N=7 for Adult Men from Matero

**N=7 for Elderly from George

**Table S3 | Probability that each HWF is ranked first overall before and after price is considered.** *Stars represent where p-values fell when comparing each item's log odds of being ranked higher compared to a reference item (ref).*

| HWF | Probability HWF Ranked First (%) | | | |
|---|---|---|---|---|
| | Overall | | Overall + Price | |
| Happy Tap | 34% | *** | 9% | *** |
| Jengu | 28% | *** | 5% | |
| Kalingalinga | 24% | *** | 44% | *** |
| Kohler | 5% | *** | 8% | ** |
| Sanitap | 2% | | 6% | |
| Satotap | 2% | * | 10% | *** |
| Tippytap | 3% | *** | 13% | *** |
| Spatap | 2% | (Ref) | 5% | (Ref) |

*P<0.05

**P<0.01

***P<0.001

**Table S4 | Probability that each HWF is ranked first overall before price is considered, stratified by group.** *Stars represent where p-values fell when comparing each item's log odds of being ranked higher compared to a reference item (ref).*

| HWF | Probability HWF Ranked First Overall (%) | | | | | | | |
|---|---|---|---|---|---|---|---|---|
| | Adult Men | | Adult Women | | Caregivers | | Elderly | |
| Happy Tap | 27% | *** | 33% | *** | 29% | *** | 35% | *** |
| Jengu | 26% | *** | 46% | *** | 41% | *** | 11% | ** |
| Kalingalinga | 36% | *** | 10% | *** | 21% | *** | 29% | *** |
| Kohler | 5% | *** | 5% | ** | 4% | *** | 4% | |
| Sanitap | 1% | | 1% | | 1% | | 4% | |
| Satotap | 2% | * | 1% | | 1% | | 5% | |
| Tippytap | 1% | | 1% | | 2% | ** | 10% | ** |
| Spatap | 1% | (Ref) | 2% | (Ref) | 1% | (Ref) | 2% | (Ref) |

*P<0.05

**P<0.01

***P<0.001

**Table S5| Probability that each HWF is ranked first overall after price is considered, stratified by group.** *Stars represent where p-values fell when comparing each item's log odds of being ranked higher compared to a reference item (ref).*

| HWF | Probability HWF Ranked First Overall + Price (%) | | | | | | | |
|---|---|---|---|---|---|---|---|---|
| | Adult Men | | Adult Women | | Caregivers | | Elderly | |
| Happy Tap | 8% | ** | 12% | | 4% | | 16% | * |
| Jengu | 5% | | 7% | | 2% | | 8% | |
| Kalingalinga | 54% | *** | 36% | *** | 61% | *** | 28% | *** |
| Kohler | 5% | * | 12% | | 4% | | 12% | |
| Sanitap | 6% | ** | 9% | | 3% | | 6% | |
| Satotap | 10% | *** | 9% | | 7% | | 10% | |
| Tippytap | 10% | *** | 9% | | 15% | *** | 14% | |
| Spatap | 2% | (Ref) | 6% | (Ref) | 3% | (Ref) | 7% | (Ref) |

*P<0.05

**P<0.01

***P<0.001

**Table S6| Probability that each HWF is ranked first overall before price is considered, stratified by community.** *Stars represent where p-values fell when comparing each item's log odds of being ranked higher compared to a reference item (ref).*

| HWF | Probability HWF Ranked First Overall (%) | | | |
|---|---|---|---|---|
| | George | | Matero | |
| Happy Tap | 38% | *** | 28% | *** |
| Jengu | 24% | *** | 33% | *** |
| Kalingalinga | 28% | *** | 18% | *** |
| Kohler | 4% | *** | 7% | *** |
| Sanitap | 1% | | 2% | |
| Satotap | 2% | | 3% | |
| Tippytap | 1% | * | 2% | ** |
| Spatap | 2% | (Ref) | 6% | (Ref) |

*P<0.05

**P<0.01

***P<0.001

**Table S7| Probability that each HWF is ranked first overall after price is considered, stratified by community.** *Stars represent where p-values fell when comparing each item's log odds of being ranked higher compared to a reference item (ref).*

| HWF | Probability HWF Ranked First Overall + Price (%) | | | |
|---|---|---|---|---|
| | George | | Matero | |
| Happy Tap | 7% | *** | 10% | |
| Jengu | 3% | | 6% | |
| Kalingalinga | 57% | *** | 33% | *** |
| Kohler | 6% | *** | 9% | |
| Sanitap | 4% | | 9% | |
| Satotap | 9% | *** | 10% | |
| Tippytap | 3% | *** | 7% | ** |
| Spatap | 10% | (Ref) | 16% | (Ref) |

*P<0.05
**P<0.01
***P<0.001

**Table S8| Probability that each HWF is ranked first overall before price is considered, stratified by participants.** *Stars represent where p-values fell when comparing each item's log odds of being ranked higher compared to a reference item (ref).*

| HWF | Probability HWF Ranked First Overall (%) | | | |
|---|---|---|---|---|
| | Returned from Phase 1 | | New Participants | |
| Happy Tap | 31% | *** | 34% | *** |
| Jengu | 32% | *** | 27% | *** |
| Kalingalinga | 28% | *** | 23% | *** |
| Kohler | 4% | *** | 6% | *** |
| Sanitap | 1% | | 2% | |
| Satotap | 1% | | 3% | |
| Tippytap | 1% | | 4% | *** |
| Spatap | 1% | (Ref) | 2% | (Ref) |

*P<0.05
**P<0.01
***P<0.001

**Table S9| Probability that each HWF is ranked first overall before price is considered, stratified by participants.** *Stars represent where p-values fell when comparing each item's log odds of being ranked higher compared to a reference item (ref).*

| HWF | Probability HWF Ranked First Overall + Price (%) | | | |
|---|---|---|---|---|
| | Returned from Phase 1 | | New Participants | |
| Happy Tap | 8% | *** | 9% | * |
| Jengu | 3% | * | 5% | |
| Kalingalinga | 65% | *** | 41% | *** |
| Kohler | 3% | * | 10% | * |
| Sanitap | 2% | | 7% | |
| Satotap | 6% | *** | 11% | ** |
| Tippytap | 12% | *** | 13% | *** |
| Spatap | 1% | (Ref) | 6% | (Ref) |

*P<0.05
**P<0.01
***P<0.001

**Table S10| Probability that each attribute is ranked as most important, stratified by participants.** *Stars represent where p-values fell when comparing each item's log odds of being ranked higher compared to a reference item (ref).*

| HWF | Probability HWF Ranked First (%) - Attributes | | | |
|---|---|---|---|---|
| | Returned from Phase 1 | | New Participants | |
| Appearance | 7% | ** | 12% | ** |
| Convenience | 8% | ** | 12% | ** |
| Ease of Use | 15% | *** | 10% | |
| Hygienic Use | 34% | *** | 17% | *** |
| Maintenance | 6% | * | 7% | |
| Price | 4% | | 9% | |
| Water Disposal | 8% | ** | 7% | |
| Water Management | 15% | *** | 19% | *** |
| Vulnerability | 2% | (Ref) | 7% | (Ref) |

*P<0.05
**P<0.01
***P<0.001

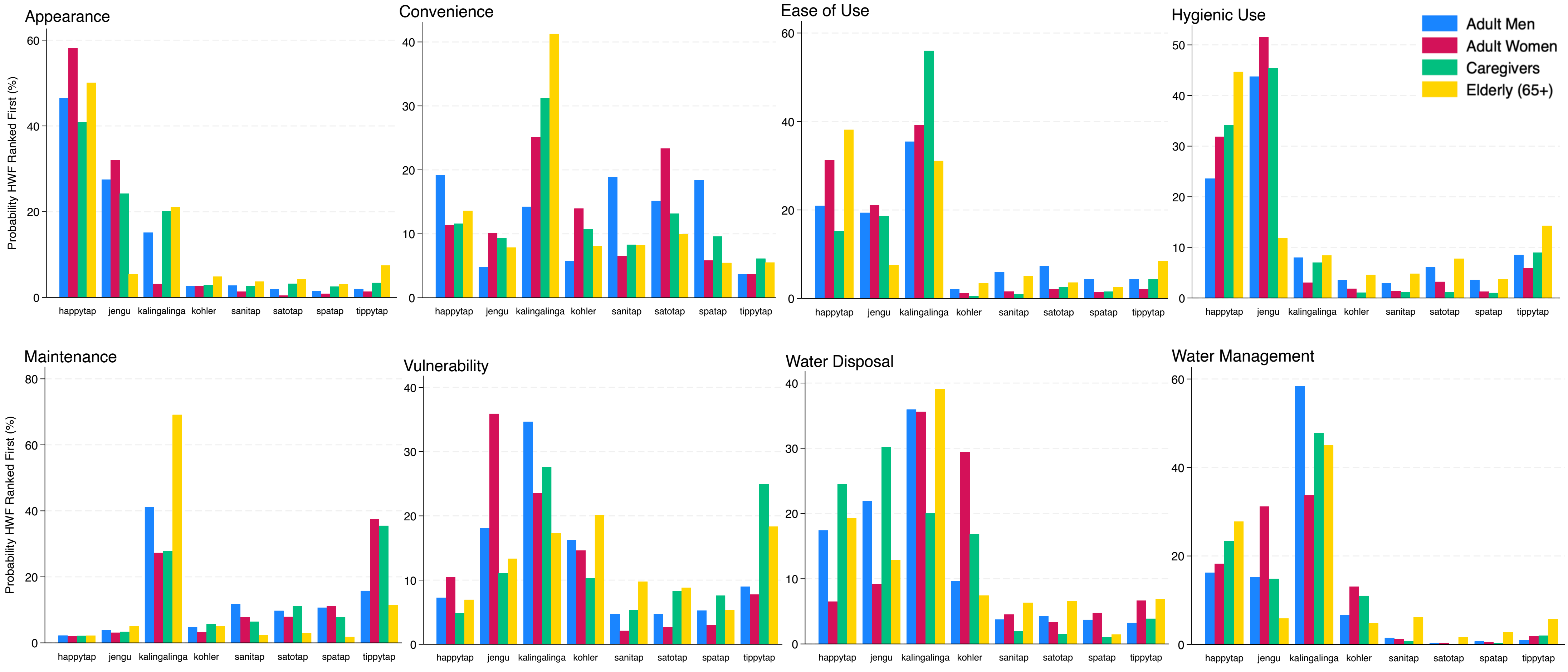

**Figure S1 | Probability of each handwashing facility being ranked first against different attributes, stratified by participant group. Predicted probabilities were estimated using rank-ordered logistic regression.**

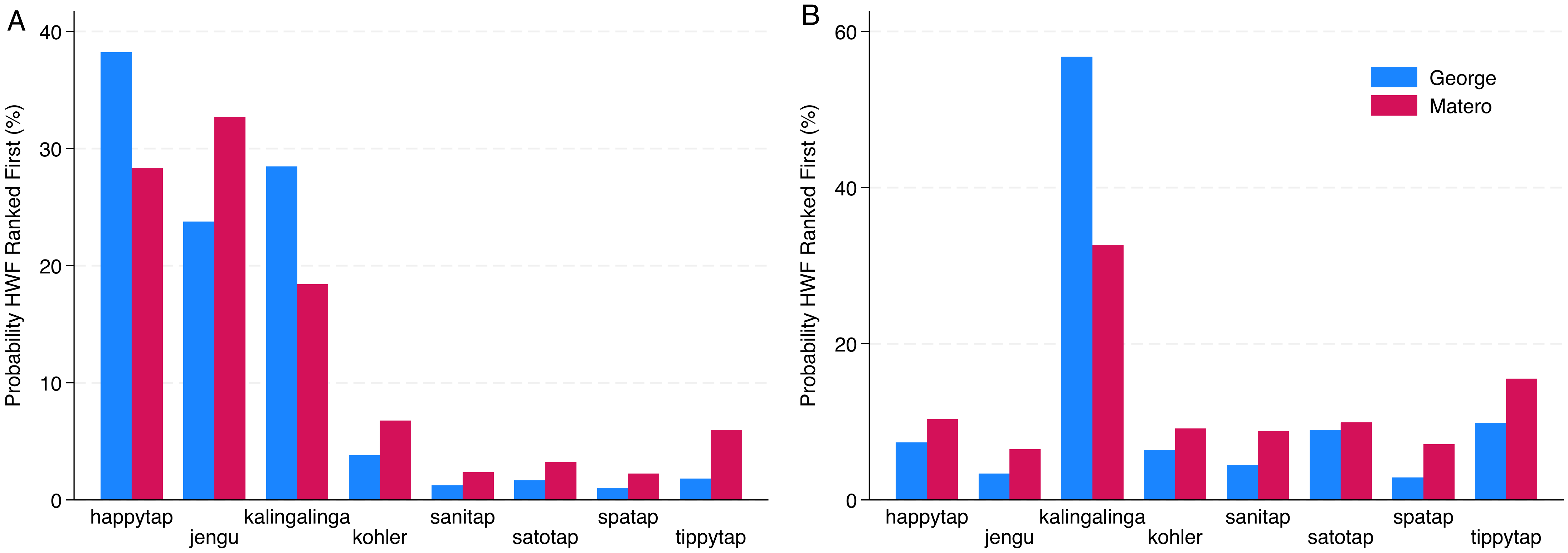

**Figure S2 | Probability of each handwashing facility being ranked first overall before (A) and after (B) consideration for price, stratified by location.** *Predicted probabilities were estimated using rank-ordered logistic regression. Wald test found no variance in overall HWF rankings between study sites before (P=0.028) and after (P=0.048) consideration for price.*
